# Plasma Cell-free RNA Signatures of Inflammatory Syndromes in Children

**DOI:** 10.1101/2024.03.06.24303645

**Authors:** Conor J. Loy, Venice Servellita, Alicia Sotomayor-Gonzalez, Andrew Bliss, Joan Lenz, Emma Belcher, Will Suslovic, Jenny Nguyen, Meagan E. Williams, Miriam Oseguera, Michael A. Gardiner, Pediatric Emergency Medicine Kawasaki Disease Research Group (PEMKDRG), The CHARMS Study Group, Jong-Ha Choi, Hui-Mien Hsiao, Hao Wang, Jihoon Kim, Chisato Shimizu, Adriana Tremoulet, Meghan Delaney, Roberta L. DeBiasi, Christina A. Rostad, Jane C. Burns, Charles Y. Chiu, Iwijn De Vlaminck

**Author notes:** Co-corresponding authors: Jane Burns, Charles Chiu, Iwijn De Vlaminck.

## Abstract

Inflammatory syndromes, including those caused by infection, are a major cause of hospital admissions among children and are often misdiagnosed because of a lack of advanced molecular diagnostic tools. In this study, we explored the utility of circulating cell-free RNA (cfRNA) in plasma as an analyte for the differential diagnosis and characterization of pediatric inflammatory syndromes. We profiled cfRNA in 370 plasma samples from pediatric patients with a range of inflammatory conditions, including Kawasaki disease (KD), Multisystem Inflammatory Syndrome in Children (MIS-C), viral infections and bacterial infections. We developed machine learning models based on these cfRNA profiles, which effectively differentiated KD from MIS-C — two conditions presenting with overlapping symptoms — with high performance (Test Area Under the Curve (AUC) = 0.97). We further extended this methodology into a multiclass machine learning framework that achieved 81% accuracy in distinguishing among KD, MIS-C, viral, and bacterial infections. We further demonstrated that cfRNA profiles can be used to quantify injury to specific tissues and organs, including the liver, heart, endothelium, nervous system, and the upper respiratory tract. Overall, this study identified cfRNA as a versatile analyte for the differential diagnosis and characterization of a wide range of pediatric inflammatory syndromes.

## INTRODUCTION

The differential diagnosis of inflammatory syndromes in children is complex owing to their overlapping clinical manifestations, non-specific symptoms and developmental age-related barriers to communication. These challenges often result in delayed or inaccurate diagnoses, thereby impeding effective clinical management and increasing the risk of long-term adverse health effects. A key example is Kawasaki disease (KD), an inflammatory syndrome of unknown etiology that primarily affects children under five years of age. KD is often misdiagnosed because significant clinical signs overlap between KD and other inflammatory and/or infectious conditions. Accurate diagnosis is critical, as KD patients who do not receive intravenous immunoglobulin (IVIG) early in the course of illness have a substantially increased risk of developing coronary artery aneurysms, making KD the leading cause of acquired heart disease in children. Thus, there is a clear need for accurate molecular tests for inflammatory conditions such as KD to inform timely and appropriate treatment.

Currently, the differential diagnosis of pediatric inflammatory syndromes relies on clinical assessment of signs and symptoms and results from a broad array of laboratory tests. Culture-based and molecular assays are routinely used to identify viral and bacterial pathogens, but these tests do not interrogate the host response and hence are unable to differentiate between infectious and non-infectious conditions. Serologic metabolic, and antigen biomarkers used for diagnosis often lack specificity^1,2^. To address these limitations, recent studies have explored the use of whole blood RNA transcriptome profiling of the human host response to assess disease severity and differentiate among inflammatory conditions including KD, MIS-C, viral, and bacterial infection^3–6^. However, while the whole blood profile is indicative of the host immune response, it provides little information regarding the extent of inflammation-related cell injury or death in solid organ tissues. In contrast, cell-free nucleic acids in plasma, including cell-free DNA (cfDNA) and cell-free RNA (cfRNA) are promising analytes for evaluating inflammation as they are released by dead or dying cells originating from both the bloodstream and solid tissues^7^. Many recent studies have explored the use of cell-free nucleic acids for monitoring of pregnancy^8,9^, cancer^10,11^, transplantation^12,13^ and infection^5,14–23^, yet the potential of cell-free nucleic acids for the differential diagnosis of inflammatory syndromes remains largely unexplored.

In this study, we applied plasma cell-free RNA (cfRNA) profiling by RNA sequencing to compare host immune and cellular injury responses associated with four different inflammatory and/or infectious syndromes in children. We identified shared signatures across multiple conditions, highlighting the importance of incorporating multiple comparison groups for the development of disease-specific biomarkers. Using cfRNA profiles, we built a machine learning model with high accuracy in differentiating between MIS-C and KD, two pediatric inflammatory syndromes with overlapping clinical presentations. We then expanded this methodology to construct a multi-class diagnostic classifier capable of distinguishing among KD, MIS-C, viral, and bacterial infections. In addition, we demonstrate that the cfRNA profile can be correlated with markers of tissue damage and may be able to differentiate among different viral infections. We propose an application of cfRNA profiling as a decision support tool for differential diagnosis of inflammatory syndromes in the clinical setting.

## RESULTS

### Clinical cohort

We collected and analyzed 370 plasma samples from pediatric patients with inflammatory and infectious conditions at four hospitals in the US, including Rad Children’s Hospital San Diego (UCSD), Emory, Children’s National Hospital (CNH), and University of California San Francisco (UCSF) (**Figure 1A and B).** This sample set included patients diagnosed with KD, MIS-C, viral infection, bacterial infection, and other hospitalized pediatric controls, some but not all with inflammatory disease (for example, arthralgia, Crohn’s disease flare, parenchymal lung disease, chronic lung disease, toxic shock syndrome, and post-vaccine myocarditis), as well as healthy children (**Figure 1A** and **Table 1**). Included cases of bacterial and viral infections were heterogeneous with respect to infection site and pathogen (**Supplementary File 1**). All patient samples were collected within 4 days of hospitalization during the acute phase of disease (Methods). RNA next-generation sequencing was used to quantify the genes and cell types of origin of cfRNA in each sample, with an average of 20.7 million reads sequenced per sample (Methods). Machine learning models were built to identify biosignatures associated with different diseases.

**Figure 1.**
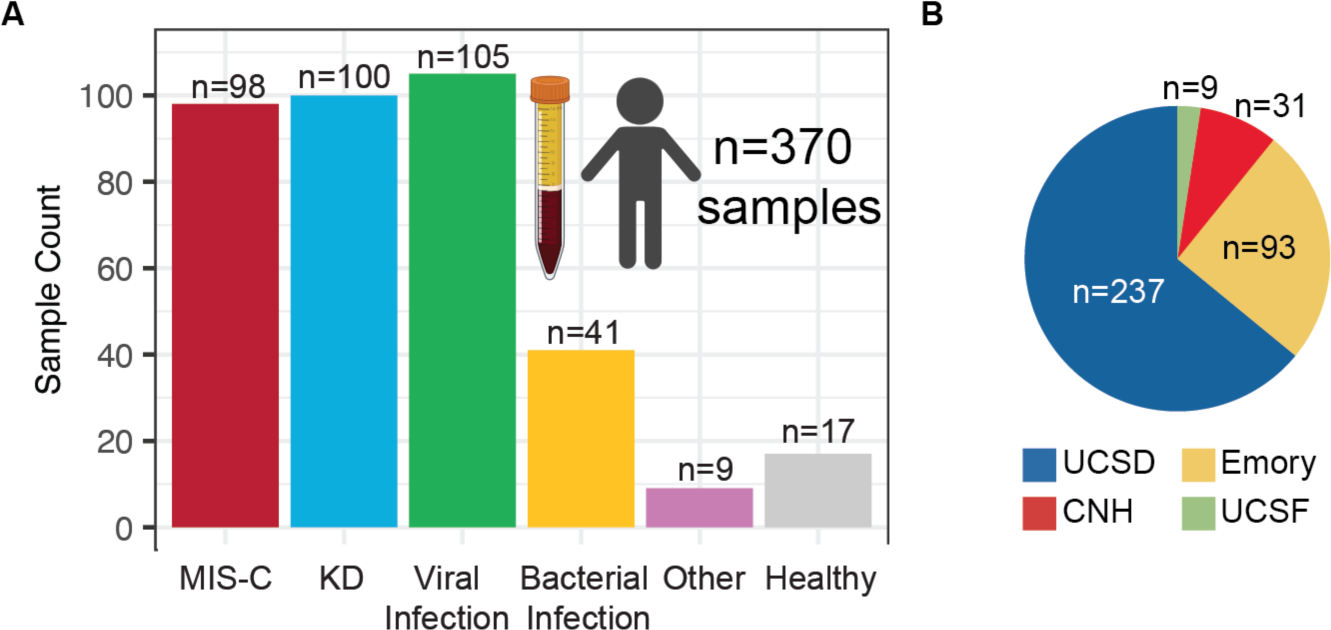
Sample Overview. **A)** Sample counts for each disease group. “Other” indicates other hospitalized pediatric controls. **B)** Distribution of collected samples based on hospital site.

### cfRNA signatures of disease

Our initial focus was on characterizing changes in cfRNA profiles in plasma that are common across the various inflammatory conditions. We performed pairwise differential gene abundance analysis between the healthy control group and each disease group (Methods, **Figure 2A** and **Supplementary File 2**). This analysis identified differentially abundant genes (DAGs) for all comparisons (BH adjusted p-value < 0.05). The smallest number of DAGs was identified when comparing healthy controls to patients in the other hospitalized control group (n=2,686) and the greatest number when comparing healthy controls to patients with KD (n=6,591, **Figure 2A**). Further analysis of the DAGs for all conditions revealed a significant number of shared genes (1,877 DAGs, **Figure 2A**). Chief among these were histone protein coding genes that were elevated for all disease groups, indicating that an increase in histone transcripts is a universal indicator of inflammation (**Figure S1A**). The analysis also identified immune related transcripts MPO, ELANE, CD53, and CXCR2 elevated in each disease group, homeostasis related transcripts CDK19, ANAPC5, and 32 mitochondrial protein coding RNAs in the control group (**Figure S2B**). Pathway analysis of the 1,877 overlapping DAGs revealed an enrichment of neutrophil and cell replication transcripts related to inflammation (**Figure 2B** and **Supplementary File 3**). These findings point to shared signatures of inflammation among disease groups and emphasize the need for inter-group comparisons to develop disease-specific biosignatures.

**Figure 2.**
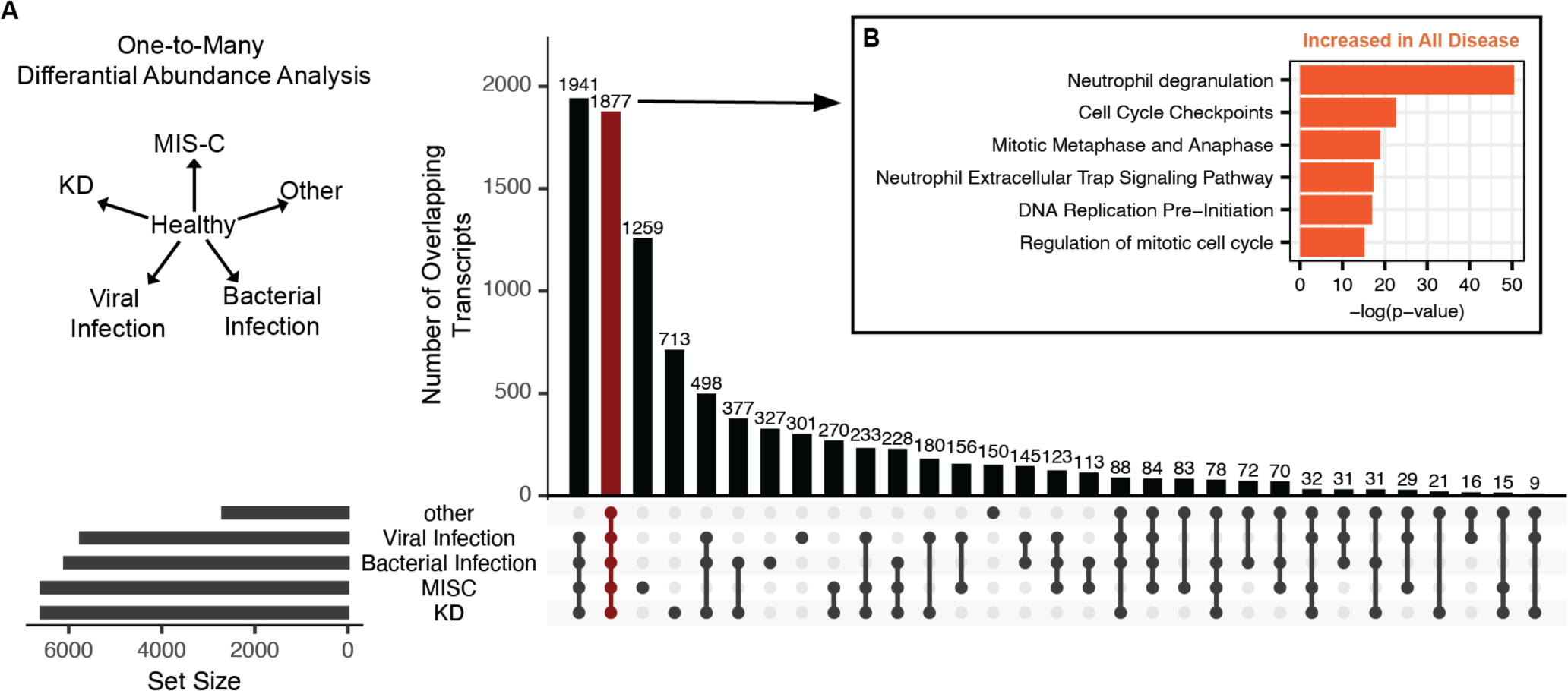
Shared cfRNA signatures among different inflammation syndromes. **A)** Differential abundance analysis using DESeq2 was performed between healthy controls and each other group individually. Vertical columns indicate the number of overlapping genes that are significantly differentially abundant between groups and controls (Benjamini-Hochberg (BH) adjusted p-values < 0.05). Dots below bars indicate the groups being intersected. Horizontal columns indicate the total number of DAGs between groups and controls. **B)** Significantly enriched pathways in the set of genes found to be differentially abundant between healthy controls and each disease group group. Average p-value and fold change used for pathway analysis (Qiagen, IPA).

### cfRNA can differentiate MIS-C and Kawasaki Disease

KD and MIS-C share many clinical characteristics: they are highly inflammatory, present with endothelial dysfunction, and have multiple overlapping signs, including skin rashes, mucosal involvement, and fever. There are currently no molecular tests to distinguish between KD and MIS-C. We therefore assessed if cfRNA could be used to differentiate these severe pediatric inflammatory syndromes. We divided the KD (n=100) and MIS-C (n=98) samples into training (60%), validation (20%), and test (20%) sets, ensuring a roughly equal representation of hospital of origin and disease subclassification across all 3 sets (**Figure 3A**). We used the training data for feature selection and to train machine learning models. We then used the validation set to select the final model, based on the model with the highest receiving operator characteristic Area Under the Curve (AUC). Last, we evaluated the performance of the final model using the test set. To prevent influence of the test set on the training data and to ensure unbiased results, we normalized each set separately using a variance stabilizing transformation (Methods).

**Figure 3.**
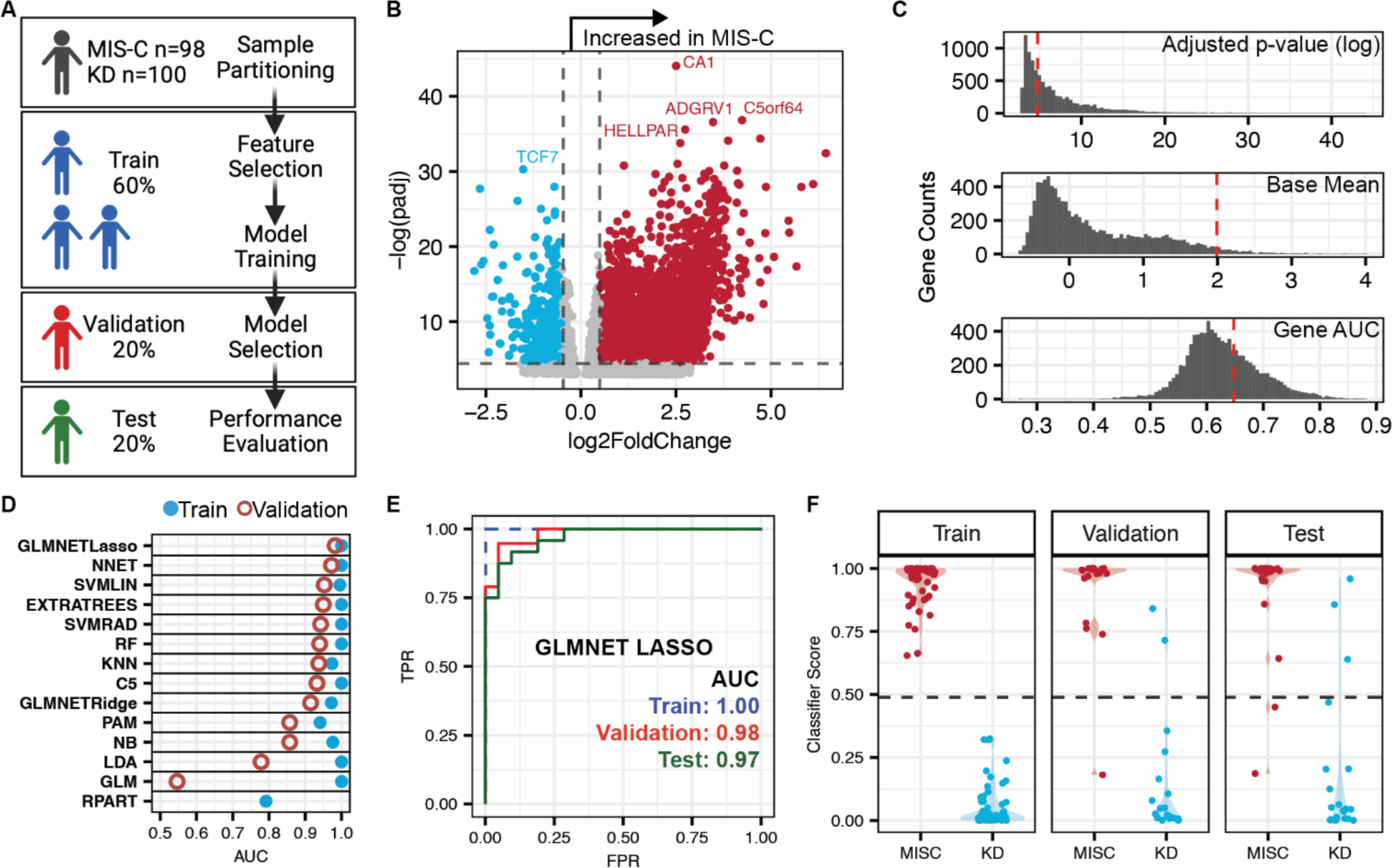
cfRNA distinguishes KD and MIS-C. **A)** Overview of sample set and modeling scheme. **B)** Volcano plot of differentially abundant transcripts between MIS-C and KD. Analysis was performed using the training data set (DESeq2). **C)** Adjusted p-value, base mean, and gene ROC AUC distributions for all genes from the training KD vs MIS-C comparison. **D)** ROC AUC values for the 14 machine learning classification models applied to training and validation sets. **E)** ROC-AUC curves of the training, validation, and test sets using the GLMNET with LASSO regression algorithm. **F)** Violin plots of the classifier scores from the GLMNET with LASSO regression algorithm.

In our initial analysis using the training set, we identified 1,242 differentially abundant genes between KD and MIS-C (DESeq2, Benjamini-Hochberg (BH) adjusted p-value < 0.01 and base mean > 10; **Figure 3B** and **Supplementary File 4**). We refined this gene list based on adjusted p-value, base mean, individual gene AUC, and fold change, resulting in a final tally of 132 genes for model input (**Figure 3C**). We then trained 14 machine learning classification models, evaluating their performance on the validation set (Methods). The GLMNET model with LASSO regression exhibited the highest validation set AUC and was selected as our final model (**Figure 3D**). A unique feature of the GLMNET LASSO algorithm is the feature selection step, which selected 25 genes for the modeling (**Table S1**). Using this trained model, we tested the classification performance using the test set (ROC-AUC train=1.00, validation=0.98, test=0.97) (**Figure 3E**). We also observed similar distributions in the classification scores across sample sets, further confirming that there was little to no overfitting of the training and test sets (**Figure 3F**).

### Multi-disease classification using cfRNA

We next asked if cfRNA could be used in the more challenging scenario of multi-disease classification. For this, we developed a machine learning framework that combines the outputs of one-vs-one models using a random forest multiclass algorithm (Methods, **Figure 4A**). We first split our dataset into training (70%) and test (30%) sets, with roughly comparable proportions of samples from patients with KD, MIS-C, viral infection, or bacterial infection, while also stratifying the groups evenly with respect to hospital of origin and disease subclassification. We performed differential abundance analysis for each pairwise comparison using only the training data (**Supplementary File 5**). Next, we trained individual one-vs-one GLMNET models with LASSO regression for each sample group (MIS-C, KD, viral infection, or bacterial infection), using the top 100 genes identified in the differential abundance analysis. These genes were selected based on gene abundance AUC and level of significance (Methods). Each of the one-vs-one models demonstrated high performance with individual model scores classifying samples with high accuracy (Test AUC: min=0.86, max=0.99; Test Accuracy: min=0.83, max=0.92; **Figure 4B**). The union of genes used by each GLMNET model with LASSO regression generated a 109-gene panel (**Supplementary File 6**). To combine the outputs of the individual models, we trained a multiclass random forest classifier using the classification scores from the one-vs-one models. Using this framework, our multiclass machine learning model achieved high accuracy in both the training and test sets (Accuracy, train=100% and test=81%), with the highest rate of misclassified samples in the bacterial infection group. The high performance of the multiclass machine learning model on a relatively small number of genes points to the potential utility of cfRNA in differentiating complex inflammatory conditions in a clinical setting.

**Figure 4.**
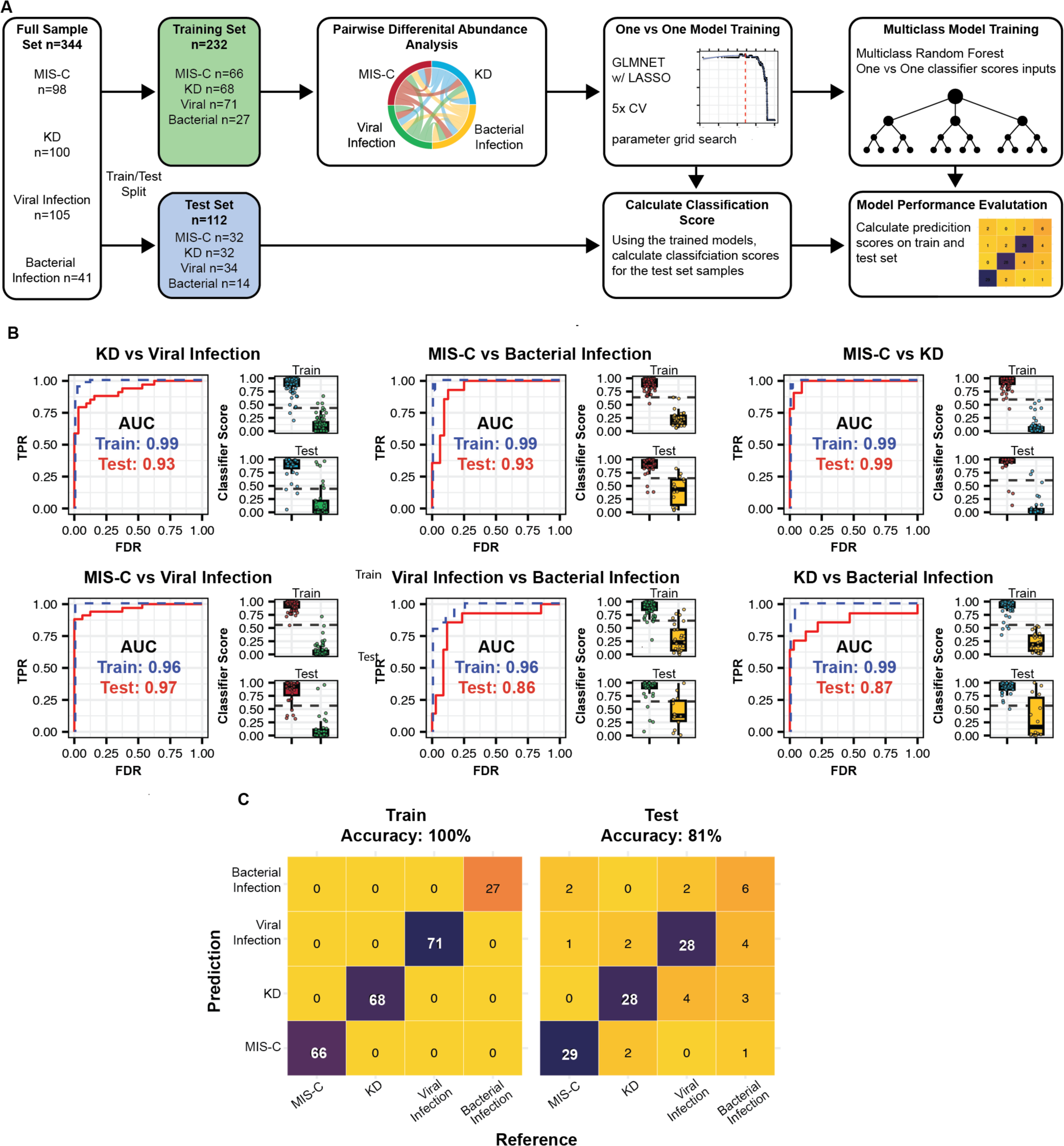
Multiclass Classification of pediatric disease using cfRNA. **A)** Overview of machine learning framework used for multiclass classification. **B)** ROC-AUC plots for each one-vs-one model trained, along with train (top) and test (bottom) classifier score distributions. **C)** Confusion matrix of reference and predicted diagnoses for train and test samples. Color indicates number of samples in each category.

### Characterization of disease using cfRNA

We further investigated whether cfRNA could be employed not only for classification but also for disease characterization. Since inflammation and/or infection can impact multiple organ systems, understanding organ-specific damage and function is crucial for guiding clinical management. Previous work from our group and others has shown that quantifying the cfRNA cell-type-of-origin (CTO) is a viable non-invasive method to assess cell, tissue and organ-specific injury^17,24^. Here, we employed deconvolution of the cell types of origin of cfRNA using BayesPrism and the Tabula Sapiens human cell atlas as a reference. The CTO estimates were compared with known biomarkers and other indicators of specific organ injury or dysfunction (Methods, **Figure 5A**). We first compared hepatocyte and intrahepatic cholangiocyte contributions to the cfRNA in plasma to alanine transaminase measurements (ALT, n=141). We observed significantly elevated levels of cfRNA from hepatocytes and intrahepatic cholangiocytes in patients with high ALT (ALT > 100 IU/L) compared to patients with normal ALT (ALT < 40 IU/L, **Figure 5A**). To explore cardiac function and damage, samples were categorized as having either normal or abnormal cardiac function (Methods). In the abnormal group, we observed increased levels of cfRNA from cardiac muscle cells and pericytes, likely indicative of increased cardiac cell injury or death. Interestingly, this group also exhibited elevated cfRNA levels from kidney epithelial cells, intrahepatic cholangiocytes, club cells and type I pneumocytes, and bronchial epithelium, suggesting other end organ damage associated with impaired cardiac function (**Figure 5A**). Of note, patients with abnormal cardiac function did not have elevated levels of smooth or skeletal muscle cell derived cfRNA (**Figure S2A**). Last, to evaluate lung function and damage we focused on samples from healthy individuals and viral infection patients with COVID-19, further stratifying the COVID-19 samples by disease severity (Methods). We observed elevated levels of cfRNA from club cells and type I pneumocytes in moderate COVID-19 cases compared to healthy individuals, with a greater increase for severe cases (**Figure 5A**). We observed similar trends in the levels of bronchial epithelium derived cfRNA; however, the difference was not statistically significant.

**Figure 5.**
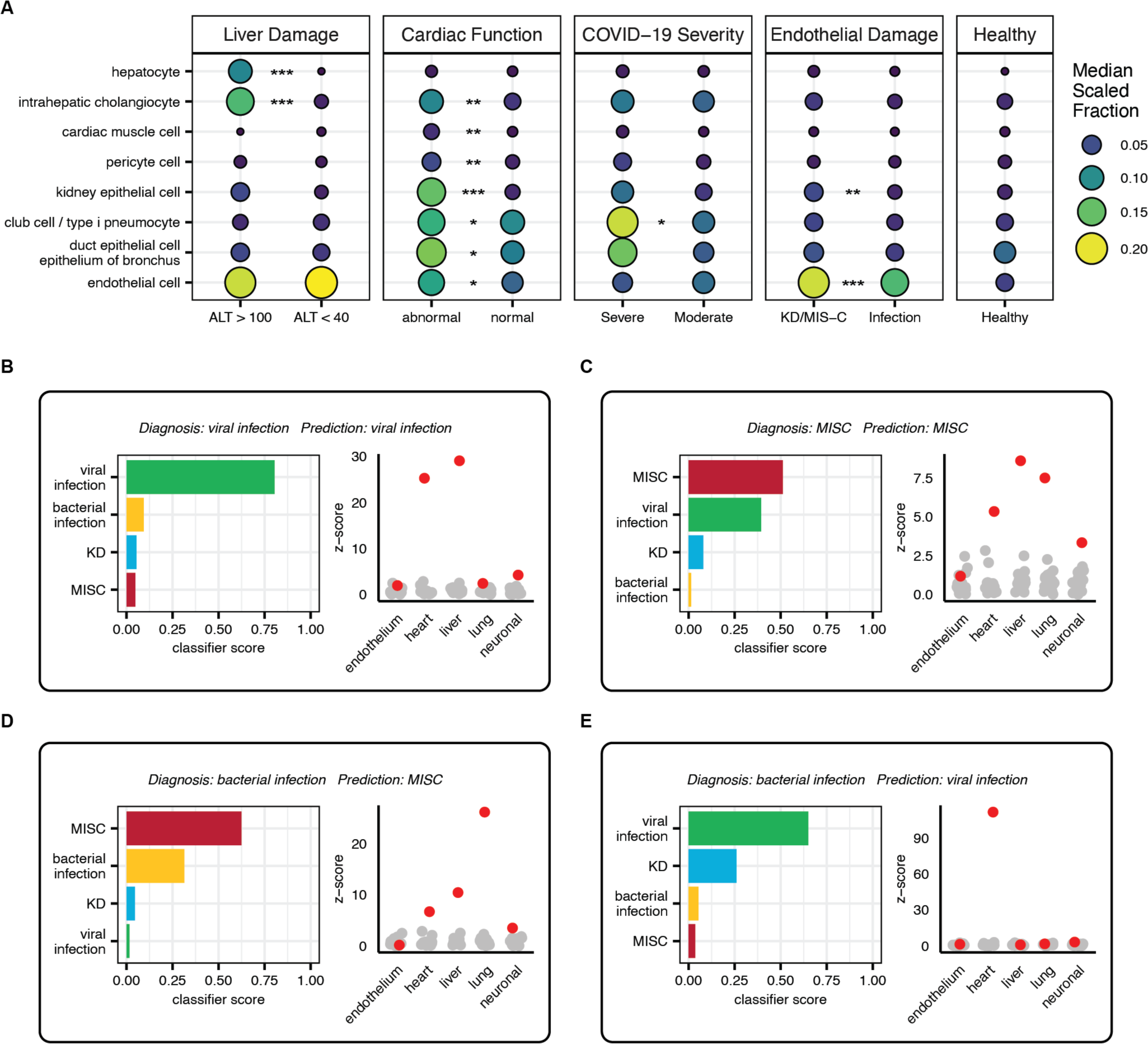
cfRNA as a clinical decision support tool. **A)** Median scaled cell type of origin fractions for samples separated by liver damage, cardiac function, COVID-19 severity, and endothelial damage, and healthy controls (Methods). Stars indicate statistically significant differences between groups in comparison (Wilcoxon rank sum test, BH adjusted p-value < 0.05). **B-E)** Case studies of patients with example clinical decision support tool results from the multiclass algorithm, along with measurements of endothelium, heart, liver, lung, and neuronal damage from the deconvolution data. Sample shown in red and healthy donor samples in gray. Z-scores calculated relative to healthy donor sample distributions. Endothelium refers to endothelial cell, heart to cardiac muscle cells, liver to hepatocyte, lung to club cell and type I pneumocyte, and neuronal to Schwann cell cfRNA cell type of origin fractions.

We next assessed if cfRNA could be used to further stratify sample groups by distinguishing between COVID-19 infection from SARS-CoV-2 and infections from other viral pathogens (Influenza, RSV, EBV, Adenovirus, etc). We separated viral infection samples into COVID-19 and non-COVID-19 viral infections and randomly split the data into training (70%) and testing (30%) data sets. Next, we trained a GLMNET with LASSO regression model to differentiate between these groups (Methods). The trained model had high performance on the training and test sets despite including only seven genes (train AUC = 0.99, test AUC = 0.93), demonstrating the potential of cfRNA for respiratory viral species differentiation (**Figure S2B-C)**.

### cfRNA profiling as a clinical decision support tool

We next simulated a cfRNA “report” for each patient in the multiclass test set (**Supplementary File 7)** and discuss four patient cases in detail to illustrate how cfRNA can be integrated to support clinical decision making. Each report includes diagnostic predictions from the multiclass classification algorithm and predicted organ involvement levels as z-scores of CTO fractions in samples from patients with disease compared to healthy patients.

The first case is a 10–15-year-old male with very-high-risk B-cell acute lymphoblastic leukemia in delayed intensification chemotherapy who presented with a 1-day history of fever (**Figure 5B**). His laboratory assessments were notable for leukopenia, anemia, thrombocytopenia, elevated C-reactive protein (CRP), mildly elevated alanine transaminase, normal kidney function, and developed hypotension following admission. He had known household contacts with COVID-19, and his nasopharyngeal PCR was positive for SARS-CoV-2, but he did not develop respiratory symptoms. The final diagnosis was COVID-19 in an immunocompromised host. Despite the immunocompromised state of the patient and lack of respiratory symptoms common in COVID-19, the cfRNA algorithm correctly identified the patient as having a viral infection. Furthermore, the elevated levels of liver and heart derived cfRNA are interesting given the clinical manifestations of mildly elevated ALT and hypotension.

In the second case, the cfRNA model correctly predicted the patient as having MIS-C, but the classifier score for MIS-C and viral infection were very comparable, producing what could be considered a “borderline” result (**Figure 5C**). The patient is a 0–5-year-old male who presented with a 1-week history of abdominal pain, vomiting, and watery diarrhea, with subsequent development of fever, conjunctival injection and swelling of the hands and feet. The patient had normal white blood cell counts (WBC), with lymphopenia, thrombocytopenia, hyponatremia, acidosis, and elevated brain natriuretic peptide (BNP), C-reactive protein (CRP) and erythrocyte sedimentation rate (ESR), with normal liver and kidney function. He was SARS-CoV-2 PCR negative but SARS-CoV-2 immunoglobulin G positive. No additional viral testing was performed. His course was complicated by hypotension, but an echocardiogram was grossly normal. The final diagnosis was MIS-C, which was the top prediction from the cfRNA model. Interestingly, viral infection was the second most probable diagnosis from the cfRNA model. MIS-C is derived from a viral infection (SARS-CoV-2), and MIS-C and viral infection can be difficult to differentiate. There are at least 3 possible interpretations for these results: (1) the model is correctly identifying MIS-C along with a viral signature from SARS-CoV-2, (2) the patient is misdiagnosed with MIS-C and has only a viral infection, or (3) the detection of a viral signature is due to errors in the model. We also observed elevated liver cfRNA despite the patient having normal liver enzymes along with elevated heart tissue derived cfRNA, which is consistent with the elevated BNP.

In the third case, the model incorrectly predicted the diagnosis in an immunocompromised patient, but the correct diagnosis of bacterial infection was the second most likely prediction (**Figure 5D**). The patient is a 0-5-year-old male with a history of Wiskott-Aldrich syndrome requiring a bone marrow transplant. He was admitted with fever, fatigue, pancytopenia with severe thrombocytopenia, elevated CRP, and blood cultures positive for *Klebsiella pneumoniae*. The source of the infection is unknown but was believed to be secondary to bacterial gut translocation or central line infection. His pancytopenia and thrombocytopenia were attributed to his history of Wiskott-Aldrich syndrome. He was treated with cefepime and responded clinically to a full course of antibiotics. The final diagnosis was bacterial infection, which was the second ranked prediction by the cfRNA model. Interestingly, the patient presented with some symptoms characteristic of MIS-C, the apparently erroneous top prediction made by the cfRNA model; specifically with severe thrombocytopenia and elevated CRP, which are part of the MIS-C case definition^25^.

In the last case, the model predicted an incorrect diagnosis, but the cfRNA organ damage scores in the patient were consistent with what was observed clinically (**Figure 5E**). The patient is a 0-5-year-old female presenting with 2 months of fever, progressive fatigue, and diffuse muscle pain. On admission, the patient was found to have peripheral leukocytosis (WBC count of 24.6) with >20% blasts, relative neutropenia and tumor lysis syndrome. The leukemia was subtyped as B-cell acute lymphocytic leukemia by flow cytometry. The patient was positive for methicillin-sensitive *Staphylococcus aureus* (MSSA) bacteremia with 1 of 1 culture positive. Hazy opacities observed on the chest x-ray raised concerns about pulmonary edema, while the echocardiogram indicated coronary dilation and cardiac hypertrophy which was attributed to chronic anemia. The patient responded to a full course of antibiotics, and the final diagnosis was MSSA bacterial infection, which the cfRNA model did not correctly predict. However, despite the incorrect diagnosis, organ damage scores clearly show elevated levels of heart derived cfRNA, consistent with the abnormal cardiac hypertrophy observed in this patient.

## DISCUSSION

In this work, we established a proof-of-concept that cfRNA can be used to differentiate among inflammatory syndromes in children across both one-on-one and multiclass comparisons. Interest in the use of blood-borne nucleic acids for diagnosis of inflammatory conditions is driven by continual advancements in sequencing technology, the ease of drawing blood, and the critical need for better diagnostic tools. Coote et al. recently introduced whole blood RNA multi-classification models for pediatric illnesses, distinguishing 6 broad and 18 granular categories. However, whole blood RNA biosignatures are primarily derived from immune cells, and do not provide information on cell, tissue, and organ damage. In contrast, cfRNA signals derive from dead and dying cells in blood and tissues, thus providing insight into immune dynamics and underlying tissue involvement^8–10,17–19,24,26,27^. Kalantar et al. used a combination of cfRNA, cfDNA, and whole blood RNA for differentiating sepsis and other causes of critical illness^5^. However, the approach used by Kalantar et al. lacks consolidation into a unified multi-class model, requires the use of two different analytes (whole blood and plasma), and did not analyze cfRNA to characterize tissue and organ injury. Our work expands on these recent studies by presenting a multiclass framework using only cfRNA to differentiate, diagnose, and better characterize inflammatory syndromes in children.

The data from this study provides opportunities to better understand cfRNA profiles in health and disease. We observed cfRNA signatures that are common to all inflammatory conditions, including elevated histone related RNAs, neutrophil extracellular trap (NET) components MPO and ELANE^28^, as well as immune markers ILF2, IFI16, CD53, and CXCR2. NETs are composed of DNA, histones, and other proteins, and act to trap and neutralize pathogens while minimizing host cell damage^29^. Our observation of cfRNA signatures of NET formation is consistent with the reported role of NETs in KD and MIS-C^30,31^.

To test the ability of cfRNA profiles to distinguish among pediatric inflammatory syndromes, we compared cfRNA profiles for patients diagnosed with KD and MIS-C, conditions that are very similar clinically. There is a need for improved molecular tools to discriminate between these two conditions as exemplified by a recent study from Day-Lewis et al. which reported significant overlap in signs and symptoms between KD and MIS-C (based on the 2023 case definition), with an estimated false positive rate of 8%^25^. The cfRNA signature presented here has near-perfect accuracy and therefore has high potential for translation into a useful clinical molecular test. Furthermore, the identified gene signature provides new mechanistic insight into KD and MIS-C. For example, two genes included in the machine learning signature are EEF2, elevated in KD, and FKBP5, elevated in MIS-C. EEF2 is an elongation factor that has previously been implicated in senescence^32^ and exposure to bacterial toxins^33^. This is compelling given that it has been suggested that an inhaled toxin or pathogen may be one of the triggers of KD. FKBP5 is a member of the immunophilin family and has been implicated in immune-stress response and other cellular processes in the brain and peripheral nervous system^34^. This observation is relevant given the neuronal involvement reported for MIS-C^35^. Beyond KD and MIS-C, we show that cfRNA can similarly differentiate between COVID-19 and other viral infections, opening the door for future studies to create more granular classification algorithms that can differentiate among pathogens at the species level based on cfRNA host profiling.

The case studies focusing on both correct and incorrect predictions made by the classification model highlight the potential utility of cfRNA profiling in the clinical setting. They also underscore the importance of inclusion of as many “real-life” cases as possible during the development of classification models to maximize the specificity of their predictions. In particular, immunocompromised patients exhibit altered immune and tissue/organ damage responses, and more data will likely need to be collected and classifications models tailored to optimize performance of cfRNA profiling in this subgroup. However, we are encouraged that our results suggest that cell, tissue, and/or organ injury based on cfRNA levels can be detected even for those cases for which the model disease classification is incorrect.

Here we present the first multiclass model for differential diagnosis of inflammatory syndromes using plasma cfRNA. The final model includes just 109 genes, suggesting that translating the model to a multiplexed qRT-PCR based platform with a rapid turnaround time of a few hours is likely practical. Furthermore, one of the most compelling aspects of cfRNA profiling is the ability to quantify the extent of organ involvement. We demonstrate this concept with simultaneous quantification of injury to multiple organ systems (liver, cardiac, lung, endothelium). Available clinical tests for evaluating tissue injury (for example, alanine aminotransferase levels in the liver) enabled us to confirm the accuracy and cfRNA-based detection of organ injury. Even more compelling is the potential clinical utility of cfRNA in quantifying injury to tissues where current tests are not available or lack adequate sensitivity, such as endothelial and neuronal injury. In the future, simultaneous diagnosis and characterization of tissue injury may be critical in informing clinicians on the optimal management and treatment of their patients with inflammatory syndromes of unknown etiology.

## METHODS

### Ethics Statement

The University of California, San Francisco (UCSF) Institutional Review Board (IRB) (#21-33403), San Francisco, CA; Emory University IRB (STUDY00000723), Atlanta, GA; Children’s National Medical Center IRB (Pro00010632), Washington, DC; and Cornell University IRB for Human Participants (2012010003), New York, NY each approved the protocols for this study. All samples and patient information were de-identified for analysis and shared with collaborating institutions. At Emory University, the IRB approved protocol was a prospective enrollment study under which parents provided consent and children assent as appropriate for age. At Children’s National Medical Center and UCSF, the IRB protocols were “no subject contact” sample biobanking protocols under which consent was not obtained and data was extracted from medical charts. At University of California, San Diego (UCSD), the IRB reviewed and approved collection and sharing of samples and data (IRB #140220). Signed consent and assent were provided by the parent(s) and pediatric patient, respectively.

### Sample Acquisition

Samples were acquired from UCSF as previously described^17^. Briefly, hospitalized pediatric patients were identified as having COVID-19 by testing positive with SARS-CoV-2 real-time PCR (RT-PCR). Residual whole blood samples were collected in EDTA lavender top tubes and diluted 1:1: in DNA/RNA shield (Zymo Research). The remaining blood was centrifuged at 2500 rpm for 15 min and the available plasma was retained. All samples were stored at −80℃ freezer until used. Samples were acquired from Emory and Children’s Healthcare of Atlanta as previously described^17^. Briefly, pediatric patients were classified as having COVID-19 by SARS-CoV-2 RT-PCR and as having MIS-C if they met the CDC case definition. Controls were healthy outpatients with no known history of COVID-19 who volunteered for specimen collection. Whole blood was collected in EDTA lavender top tubes and aliquoted for plasma extraction via centrifugation at 2500 rpm for 15 min. Samples were stored at −80°C and shipped on dry ice to either UCSF or Cornell for analysis. Samples were acquired from UCSD prior to any treatment in all subjects in EDTA lavender top tubes and centrifuged at 2,000g for 10 minutes. Plasma was collected and stored at −80F. Samples were acquired from Children’s National Hospital as previously described^17^. Briefly, pediatric patients were classified as having MIS-C if they met the CDC case definition. Whole blood samples were collected and centrifuged at 1300 xG for 5 minutes at room temperature. Plasma was aliquoted into a cryovial and frozen at −80°C.

### Clinical Data

Patients were stratified as previously described^17^. For the purposes of this study, patients were classified as having MIS-C by multidisciplinary teams that adjudicated whether a patient met the CDC case definition of MIS-C. COVID-19 was defined as any patient with PCR-confirmed SARS-CoV-2 infection within the preceding 14 days who did not also meet the MIS-C case definition. Kawasaki Disease patients at UCSD met the AHA definition for complete or incomplete KD. Viral and bacterial infection patients enrolled at UCSD were adjudicated and a final diagnosis assigned by the research team (one ED clinician and one pediatric infectious disease expert) 2-3 months after the acute illness to allow time for serologies, recurrence, and clinical recovery to be assessed. Patients with a self-limited illness that resolved without treatment and for whom viral studies were either negative or not done were classified as having a “viral syndrome”. Clinical data was abstracted from the medical record and submitted into a REDCap databases housed at UCSF or UCSD.

### Sample processing and sequencing

Samples were processed as described previously^17^. Briefly, samples were received on dry ice, RNA was extracted, and libraries prepared and sequenced on a NextSeq or NovaSeq Illumina sequencer. Reads were trimmed to 61 bp and sequencing data was processed using a custom bioinformatics pipeline which included quality filtering and trimming, alignment to the human GRCh38 reference genome, and counting of gene features.

### Sample quality filtering

Using the sequencing data, quality control was performed by analyzing DNA contamination, rRNA contamination, total counts, and RNA degradation. DNA contamination was estimated by calculating the ratio of reads mapping to introns and exons. rRNA contamination was measured using SAMtools (v1.14). Total counts were calculated using featureCounts^30^ (v2.0.0). Degradation was estimated by calculating the 5’-3’ bias using Qualimap^31^ (v2.2.1). Samples were removed from analysis if either the intron to exon ratio was greater than 3, if a sample had less than 75,000 total feature counts, or if the 5’-3’ read alignment ration bias was greater than 2.

### Differential abundance analysis

Gene transcript abundances were compared using a negative binomial model implemented using the DESeq2 R package^36^. Gene transcript base mean abundance, adjusted p-value, and log2 fold change were taken from the DESeq2 Results output. Gene transcript AUC was calculated using VST transformed counts and the pROC R package^37^.

### Sample partitioning

Samples were partitioned for machine learning applications taking into consideration diagnosis, hospital of origin, and disease subclassification. MIS-C and COVID-19 samples were subclassified by severity, as previously defined^17^. KD samples were subclassified by phenotypic subclusters, as previously defined^38^. Non-COVID-19 Viral and bacterial infection samples were evenly partitioned by diagnosis and hospital of origin only.

### Machine learning: MIS-C vs KD

Samples were partitioned into training, validation, and test sets at a ratio of 60:20:20, partitioning evenly based on factors such as diagnosis, severity/subgroup, and hospital of origin. Subsequently, differential abundance analysis was conducted on the training data. Genes were filtered based on specific criteria (adjusted p-value < 0.01, base mean abundance > 100, gene transcript AUC > 0.65, and absolute log2 fold change > 0.25) and the top 150 genes, as ordered by gene transcript AUC, were selected as inputs for machine learning analyses.

Raw counts for the training, validation, and test sets were individually normalized using variance stabilizing transformation, and subsets were created based on the chosen transcript features. Fourteen machine learning classification algorithms were employed using the R package Caret (10.18637/jss.v028.i05), including generalized linear models with Ridge and LASSO feature selection (GLMNETRIDGE and GLMNETLASSO), support vector machines with linear and radial basis function kernel (SVMLin and SVMRAD), random forest (RF), random forest ExtraTrees (EXTRATREES), neural networks (NNET), linear discriminant analysis (LDA), nearest shrunken centroids (PAM), C5.0 (C5), k-nearest neighbors (KNN), naive bayes (NB), CART (RPART), and generalized linear models (GLM). Training was performed using 5-fold cross-validation and grid search hyperparameter tuning.

For each model, classification score thresholds were determined using Youden’s index on the training data. The trained models were then employed to predict labels for the validation set, and performance was assessed using the area under the receiver operating characteristic curve area under the curve (ROC-AUC). The model achieving the highest AUC on the validation set was selected as the final model and subsequently applied to the test set, which had not been utilized in any phase of model training or selection. Prediction on both the validation and test sets utilized the Youden’s index threshold derived from the training set.

### Machine learning: Multi-Classification

Samples were partitioned into train and test sets at a ratio of 70:30, considering factors such as diagnosis, severity/subgroup, and hospital of origin. One-vs-one GLMNET LASSO models were trained for each pairwise comparison of samples groups (eg. KD vs Viral Infection) using the top 150 significant features (adjusted p-value <0.05, base mean abundance > 50, and absolute log2 fold change > 1) ordered by gene transcript AUC calculated using the training data. Trained models were used to calculate classification scores for all samples in the train and test data set, resulting in six classification scores for each sample. Classification scores were used to train a multiclass Random Forest algorithm which assigned probability scores for each condition. The final condition with the highest probability score from the Random Forest was assigned as the prediction for each sample.

### CTO analysis

Cell type deconvolution was performed using BayesPrism (v1.1)^39^ with the Tabula Sapiens single-cell RNA-seq atlas (Release 1)^27^ as a reference. Cells from the Tabula Sapiens atlas were grouped as previously described in Vorperian et al.^24^. Cell types with more than 100,000 unique molecular identifiers (UMIs) were included in the reference and subsampled to 300 cells using ScanPy (v1.8.1)^40^. Deconvolution values were scaled from 0-1 for each cell type and medians calculated for plotting.

For liver damage assessment, samples were separated as having normal or high ALT measurements (normal: ALT < 40 IU/L, high: ALT > 100 IU/L). The ALT measurements were taken from the same blood draw as the plasma for cfRNA processing. For cardiac function, patients were categorized as abnormal if they had abnormal EKG/ECG and/or echocardiogram results. EKG/ECG and echocardiogram results were categorized as abnormal in the context of the patient narrative and final interpretation of the studies. For endothelial damage, samples were separated as either having KD/MIS-C or bacterial/viral infection. COVID-19 patients were determined to have moderate or severe disease using the following criteria:

#### Moderate

The patient must have been hospitalized due to COVID-19 respiratory disease and/or any systemic/non-respiratory symptoms attributed to COVID-19 (e.g., neonatal fever, dehydration, new diagnosis diabetes, acute appendicitis, necrosis of extremities, diarrhea, encephalopathy, renal insufficiency, mild coagulation abnormalities, etc.).

#### Severe

The patient must have been hospitalized for COVID-19 with either high-flow oxygen requirement (high-flow nasal cannula (NC), continuous positive airway pressure (CPAP), bilevel positive airway pressure (BIPAP), intubation with mechanical ventilation, or extracorporeal membranous oxygenation (ECMO)) and/or evidence of end-organ failure (acute renal failure requiring dialysis, coagulation abnormalities resulting in bleeding or stroke, diabetic ketoacidosis (DKA), hemodynamic instability requiring vasopressors) and/or dying from COVID-19. These patients were almost always admitted to the ICU.

### Machine learning: COVID-19 vs non-COVID19 viral infection

Training was done using the same method as the KD vs MIS-C model. Briefly, viral infection samples were partitioned into train and test sets at a ratio of 70:30, considering factors such as COVID-19 status, severity/subgroup, and hospital of origin. Subsequently, differential abundance analysis was conducted on the training data. Genes were filtered based on specific criteria (adjusted p-value < 0.01, base mean abundance > 100, gene transcript AUC > 0.65, and absolute log2 fold change > 0.25) and the top 150 genes, as ordered by gene transcript AUC, were selected as inputs for machine learning analysis.

Raw counts for the train and test sets were individually normalized using variance stabilizing transformation, and subsets were created based on the chosen transcript features. A GLMNET with LASSO regression was trained on the training set using 5-fold cross-validation and grid search hyperparameter tuning. Classification score thresholds were determined using Youden’s index on the training data. The trained models were then employed to predict labels for the test set using the Youden’s index threshold derived from the training set.

### Quantification and statistical analyses

The programming language R (v4.1.0) was utilized for all statistical analyses. Statistical significance was assessed through two-sided Wilcoxon signed-rank tests and Mann-Whitney U tests, unless specified otherwise. Machine learning algorithms were trained using the Caret R package and pipelines were run using the Snakemake workflow management system^41,42^. In boxplots, boxes denote the 25th and 75th percentiles, the band within the box signifies the median, and whiskers extend to 1.5 times the interquartile range of the hinge. The alignment of all sequencing data was performed against the GRCh38 Gencode v38 Primary Assembly, with feature counting conducted using the GRCh38 Gencode v38 Primary Assembly Annotation^43^.

## DATA AVAILABILITY

Raw sequencing data in this study cannot be deposited in a public repository due to patient privacy concerns and lack of consent for a subset of the patient samples. Instead, de-identified RNA-seq count matrices have been uploaded to the NCBI (National Center for Biotechnology Information) GEO (Gene Expression Omnibus) database and will be publicly available upon publication (GSE255555). All code has been deposited on GitHub and will be available upon publication.

## AUTHOR CONTRIBUTIONS

C.J.L., R.L.D, C.A.R, J.C.B, C.Y.C, and I.D.V conceived and designed the study. C.J.L. A.B.,,J.L., and E.B. performed sequencing experiments. W.S, J.N, M.E.W, M.O, M.A.G, J.C, H.H, C.S, A.T, M.D, R.L.D, C.A.R, J.C.B, C.Y.C, PEMKDRG, and the CHARMS Study Group identified and collected patient samples and clinical metadata. V.S, A.S.G, A.B, W.S, H.W, and J.K provided input for data analysis. C.J.L., A.B., R.L.D., C.A.R., J.C.B, C.Y.C., and I.D.V. analyzed sequencing data. M.D., R.L.D., C.A.R., J.C.B, C.Y.C., and I.D.V. supervised the study. C.J.L., C.YC., and I.D.V. wrote the manuscript and prepared the figures. All authors read and edited the manuscript and agreed to its contents.

## Data Availability

De-identified RNA-seq count matrices have been uploaded to the NCBI (National Center for Biotechnology Information) GEO (Gene Expression Omnibus) database and will be publicly available upon publication (GSE255555). All code has been deposited on GitHub and will be available upon publication.

## ACKNOWLEDGEMENTS

We would like to acknowledge staff members at the UCSF Clinical Laboratories and the UCSF Clinical Microbiology Laboratories for their help in identifying and retrieving patient whole blood samples. We thank the Cornell Genomics Center and the UCSF Center for Advanced Technology for help with sequencing libraries. At Emory, we thank Nadine Baida, Amrita Banerjee, Julia Bartol, Kushmita Bhakta, Caroline Ciric, Khalel De Castro, Khadijah Francois, Theda Gibson, Laila Hussaini, Grace Li, Wensheng Li, Austin Lu, Lisa Macoy, Clair Martin, Molly Morrison, Amy Muchinsky, Heather Nurse, Maria A. Perez, Etza Peters, Brianna Rice, Anna Siaw-Anim, Kathy Stephens, Elizabeth Grace Taylor, Ashley Tippett, Katie Zaks, and the Children’s Healthcare of Atlanta Research Laboratory for their contributions to specimen and data collection. At UCSD we thank the members of the PEMKDRG: Lukas Austin-Page, MD, Amy Bryl, MD, Joelle Donofrio-Ödmann, MD, Atim Ekpenyong, MD, David Gutglass, MD, Scott Herskovitz, MD, Paul Ishimine, MD, John Kanegaye, MD, Margaret Nguyen, MD, Mylinh Nguyen, MD, Kristy Schwartz, MD, Stacey Ulrich, MD, Tatyana Vayngortin, MD, and Elise Zimmerman, MD. We thank the members of the CHARMS study that contributed samples: Jocelyn Ang, MD, Margalit Rosenkranz, MD, Joseph Bochini, MD, Michelle Sykes, MD, Lerraughn Morgan, MD, Laura D’Addese, MD, and Maria Pilar Gutierrez, MD. We thank the patients and their families for their help to further our understanding of pediatric inflammatory conditions. This work was funded by National Institutes of Health (NIH) / National Institute of Child Health and Human Development (NICHD) grants R61HD105618, R33HD105618, and R33HD105593 (R.D., C.A.R., I.D.V., JCB, AHT, CS, and C.Y.C.). The funder had no role in study design, data collection and analysis, decision to publish, or preparation of the manuscript.

